# Multivariate Analysis of Factors Affecting COVID-19 Case and Death Rate in U.S. Counties: The Significant Effects of Black Race and Temperature

**DOI:** 10.1101/2020.04.17.20069708

**Authors:** Adam Y. Li, Theodore C Hannah, John R Durbin, Nickolas Dreher, Fiona M McAuley, Naoum Fares Marayati, Zachary Spiera, Muhammad Ali, Alex Gometz, JT Kostman, Tanvir F. Choudhri

## Abstract

**Objectives:** Coronavirus disease-19 (COVID-19) has spread rapidly around the world, and many risk factors including patient demographics, social determinants of health, environmental variables, underlying health conditions, and adherence to social distancing have been hypothesized to affect case and death rates. However, little has been done to account for the potential confounding effects of these factors. Using a large multivariate analysis, this study illuminates modulators of COVID-19 incidence and mortality in U.S. counties while controlling for risk factors across multiple domains.

**Methods:** Data on COVID-19 and various risk factors in all U.S. counties was collected from publicly available data sources through April 14, 2020. Counties with at least 50 COVID-19 cases were included in case analyses and those with at least 10 deaths were included in mortality models. The 661 counties meeting inclusion criteria for number of cases were grouped into quartiles and comparisons of risk factors were made using t-tests between the highest and lowest quartiles. Similar comparisons for 217 counties were made for above average and below average deaths/100,000. Adjusted linear and logistic regression analyses were performed to evaluate the independent effects of factors that significantly impacted cases and deaths.

**Results:** Univariate analyses demonstrated numerous significant differences between cohorts for both cases and deaths. Risk factors associated with increased cases and/or deaths per 100,000 included increased GDP per capita, decreased social distancing, increased age, increased percent Black, decreased percent Hispanic, decreased percent Asian, decreased health, increased poverty, increased diabetes, increased coronary heart disease, increased physical inactivity, increased alcohol consumption, increased tobacco use, and decreased access to primary care. Multivariate regression analyses demonstrated Black race is a risk factor for worse COVID-19 outcome independent of comorbidities, poverty, access to health care, and other mitigating factors. Lower daily temperatures was also an independent risk factor in case load but not deaths.

**Conclusions:** U.S. counties with a higher proportion of Black residents are associated with increased COVID-19 cases and deaths. However, the various suggested mechanisms, such as socioeconomic and healthcare predispositions, did not appear to drive the effect of race in our model. Counties with higher average daily temperatures are also associated with decreased COVID-19 cases but not deaths. Several theories are posited to explain these findings, including prevalence of vitamin D deficiency. Additional studies are needed to further understand these effects.

## Introduction

The first reported case of severe acute respiratory syndrome coronavirus 2 (SARS-CoV-2) causing coronavirus disease 2019 (COVID-19) was reported in Wuhan, China in December 2019.^1^ By January 2020, the virus was isolated and sequenced, and COVID-19 was reported a public health emergency by the World health Organization (WHO).^2^ Since then, COVID-19 has spread rapidly around the world with confirmed person to person transmission, an estimated R0 between 1.4-6.47, and a doubling time of 1.8 days.^3^ As of April 14, 2020, the Johns Hopkins University Coronavirus Resource Center has reported over 2 million confirmed global cases and over 0.6 million cases in the United States. Developing new therapies, identifying risk factors, and minimizing spread through social distancing remain top priorities in the fight against COVID-19.^4,5^

Previous studies have examined the effects of various risk factors on spread of COVID-19 including patient demographics,^6,7^ social determinants of health,^8^ environmental variables,^9-15^ housing,^16,17^ and underlying health conditions.^18-20^ Identifying risk factors allows public health officials to determine populations at greater risk and development targeted public health interventions. Ultimately, this may help “flatten the curve” of cases and avoid overwhelming the health care system. However, prior analyses of these risk factors have not robustly accounted for potential confounding effects.

In this study, we analyze data for COVID-19 in United States counties and ground them in the larger context of patient demographics, underlying health conditions, social determinants of health, environmental variables, and social distancing adherence. We illuminate various factors that affect COVID-19 cases and deaths, and reassess these variables while controlling for possible confounding variables in multivariate logistic and linear regressions.

## Methods

### COVID-19 Cases and Deaths Data

COVID-19 confirmed case number and death number through April 14, 2020 were obtained for each U.S. county from the Center for Systems Science and Engineering (CSSE) Coronavirus Resource Center at Johns Hopkins University. This data source contains COVID-19 case number and deaths data from all 3,143 U.S. counties in 50 states and the District of Columbia (D.C.). Cases per 100,000 people and deaths per 100,000 people for each U.S. county were calculated using CSSE and census data. Counties were excluded from the analyses if they had fewer than 50 cases or their first case occurred fewer than 3 weeks prior to the end of the study. With these constraints, 661 counties from 50 states and Washington D.C were included in case analysis. Counties were excluded from death analyses if they reported fewer than 10 deaths or the first death had occurred fewer than 2 weeks prior to the end of the study. The deaths analyses include 217 counties from 37 states and D.C. (**Table 1**).

**Table 1:** Counties Per State Included in COVID-19 Case and Death Analysis

| State | # of Counties In Case Analysis | # of Counties Death Analysis | State | # of Counties In Case Analysis | # of Counties Death Analysis |
| --- | --- | --- | --- | --- | --- |
| Alabama | 15 | 4 | Montana | 2 | 0 |
| Alaska | 2 | 0 | Nebraska | 3 | 0 |
| Arizona | 7 | 3 | Nevada | 2 | 2 |
| Arkansas | 8 | 1 | New Hampshire | 4 | 0 |
| California | 29 | 15 | New Jersey | 21 | 16 |
| Colorado | 17 | 7 | New Mexico | 6 | 1 |
| Connecticut | 8 | 6 | New York | 31 | 12 |
| Delaware | 3 | 2 | North Carolina | 27 | 2 |
| District of Columbia | 1 | 1 | North Dakota | 2 | 0 |
| Florida | 35 | 11 | Ohio | 27 | 10 |
| Georgia | 44 | 12 | Oklahoma | 9 | 3 |
| Hawaii | 2 | 0 | Oregon | 6 | 1 |
| Idaho | 4 | 0 | Pennsylvania | 31 | 13 |
| Illinois | 15 | 5 | Rhode Island | 4 | 0 |
| Indiana | 30 | 8 | South Carolina | 17 | 2 |
| Iowa | 8 | 1 | South Dakota | 2 | 0 |
| Kansas | 5 | 2 | Tennessee | 14 | 4 |
| Kentucky | 6 | 1 | Texas | 32 | 10 |
| Louisiana | 34 | 19 | Utah | 6 | 0 |
| Maine | 3 | 1 | Vermont | 4 | 1 |
| Maryland | 15 | 8 | Virginia | 23 | 2 |
| Massachusetts | 12 | 11 | Washington | 14 | 8 |
| Michigan | 23 | 6 | West Virginia | 4 | 0 |
| Minnesota | 6 | 1 | Wisconsin | 10 | 2 |
| Mississippi | 16 | 0 | Wyoming | 2 | 0 |
| Missouri | 10 | 3 |  |  |  |

### Data Sources for Covariates

Race demographics for counties was obtained from the County Health Rankings and Roadmaps Program database. Daily temperature data for counties was obtained from the National Oceanic and Atmospheric Administration. County temperature was calculated using mean temperature for a period starting 10 days before the first confirmed county case and through the most current date (April 14, 2020). Unacast social distancing data was obtained through a research agreement with the company. Data for other potential risk factors used in analysis was obtained through publicly available data sources. (**Table 2**).

**Table 2:** Publicly Available Data Sources Used In Analysis

| County Data | Source |
| --- | --- |
| COVID-19 Cases+Deaths | Center for Systems Science and Engineering (CSSE) Coronavirus Resource Center at Johns Hopkins University. (https://coronavirus.jhu.edu/map.html)<sup>21</sup> |
| COVID-19 Tests | The COVID Tracking Project (https://covidtracking.com/)<sup>28</sup> |
| 2010 Population, Population Density, Housing Density | United States Census Bureau (https://www.census.gov/en.html)<sup>26</sup> |
| 2018 Gross Domestic Product | Bureau of Economic Analysis County Data (https://www.bea.gov/data/by-place-county-m)<sup>25</sup> |
| Social Distancing: % Change in Average Mobility and Non-Essential Visits | Unacast Social Distancing Scoreboard (https://www.unacast.com/covid19/social-distancing-scoreboard)<sup>24</sup> |
| 2012-2018 Population Demographics, Health, and Social Determinants of Health Statistics | County Health Rankings and Roadmaps Program (https://www.countyhealthrankings.org/)<sup>22</sup> |
| Temperature | National Oceanic and Atmospheric Administration (https://www.ncdc.noaa.gov/)<sup>23</sup> |
| Underlying Health Conditions 2014-2016: Diabetes, Hypertension, Coronary Artery Disease, Obesity, Poverty, Pollution | United States Census Bureau American Factfinder<sup>26</sup><br>United States Diabetes Surveillance System (https://gis.cdc.gov/grasp/diabetes/DiabetesAtlas.html#)<sup>30</sup> |
| 2014 Respiratory Mortality | Institute for Health Metrics and Evaluation (http://ghdx.healthdata.org/record/ihme-data/united-states-chronic-respiratory-disease-mortality-rates-county-1980-2014)<sup>29</sup> |
| 1998-2018 Liver Mortality | Multiple Cause of Death Database (https://wonder.cdc.gov/mcd-icd10.html)<sup>27</sup> |

### Statistical Analysis

T-tests were used to compare differences between covariates for the highest quartile and lowest quartile cases per 100,000 and the top 50% versus the bottom 50% deaths per 100,000. Sequential regression modelling was used to demonstrate the effect of race and temperature on cases and deaths per 100,000. Logistic regression was used to compare counties in the highest quartile for cases per 100,000 to those in the lowest. There were an insufficient number of counties meeting the deaths per 100,000 requirements to run logistic regression, thus linear regression was used for deaths per 100,000 analyses. Results of linear regression for cases per 100,000 including all 661 counties are provided for direct comparison between cases and deaths.

The sequential model had four parts. Each model included all variables from the previous models. **Model 1** was a univariate analysis. **Model 2** added the following county macroeconomic and COVID-19-specific variables: population density, GDP/capita, COVID-19 tests per 100,000 people (state level data), COVID-19 cases/100,000 people (deaths analysis only), a marker for which State the county is in, average percent reduction in cellphone movement from day of first confirmed case to end of study, the percent of population living in overcrowded housing, and the number of days since the first confirmed COVID-19 case. **Model 3** added these county demographics and environmental variables to Model 2: percent of population over 65, proportion of Black residents, percent of the population that is female, percent of population living in rural areas, the Food Environment Index (a measure of accessibility and affordability of healthy food), the rate of violent crime per 100,000 people, the average temperature from 10 days before the first case to the end of the study, air quality measured as the average annual ambient concentrations of PM2.5, percent of the population considered to be in fair or poor health, and the poverty rate. **Model 4** was the full model and included medical comorbidities and access to health care variables in addition to all variables in Model 3. Specifically these new variables were: diabetes, obesity, physical inactivity, excessive drinking, and smoking were all reported as percent of the population; liver disease, hypertension, coronary heart disease, and chronic respiratory disease were reported as mortality per 100,000 people; and the patient to primary care physician ratio, the percent of people sleeping fewer than seven hours per night, the percent of the population without health insurance and the percent of the population who received the flu vaccine were also included

All statistical analyses were performed with Prism 8.0 (GraphPad Software, San Diego, CA). For all analyses, α = 0.05.

## Results

Our study contained data from 3,143 U.S. counties across all 50 states and the District of Columbia (D.C). 661 U.S. counties (21.0%) from 50 states and D.C. were included in case analysis (**Figure 1A**). 217 U.S. counties (6.9%) from 37 states and D.C. were included in death analysis (**Figure 1B**).

**Figure 1:**
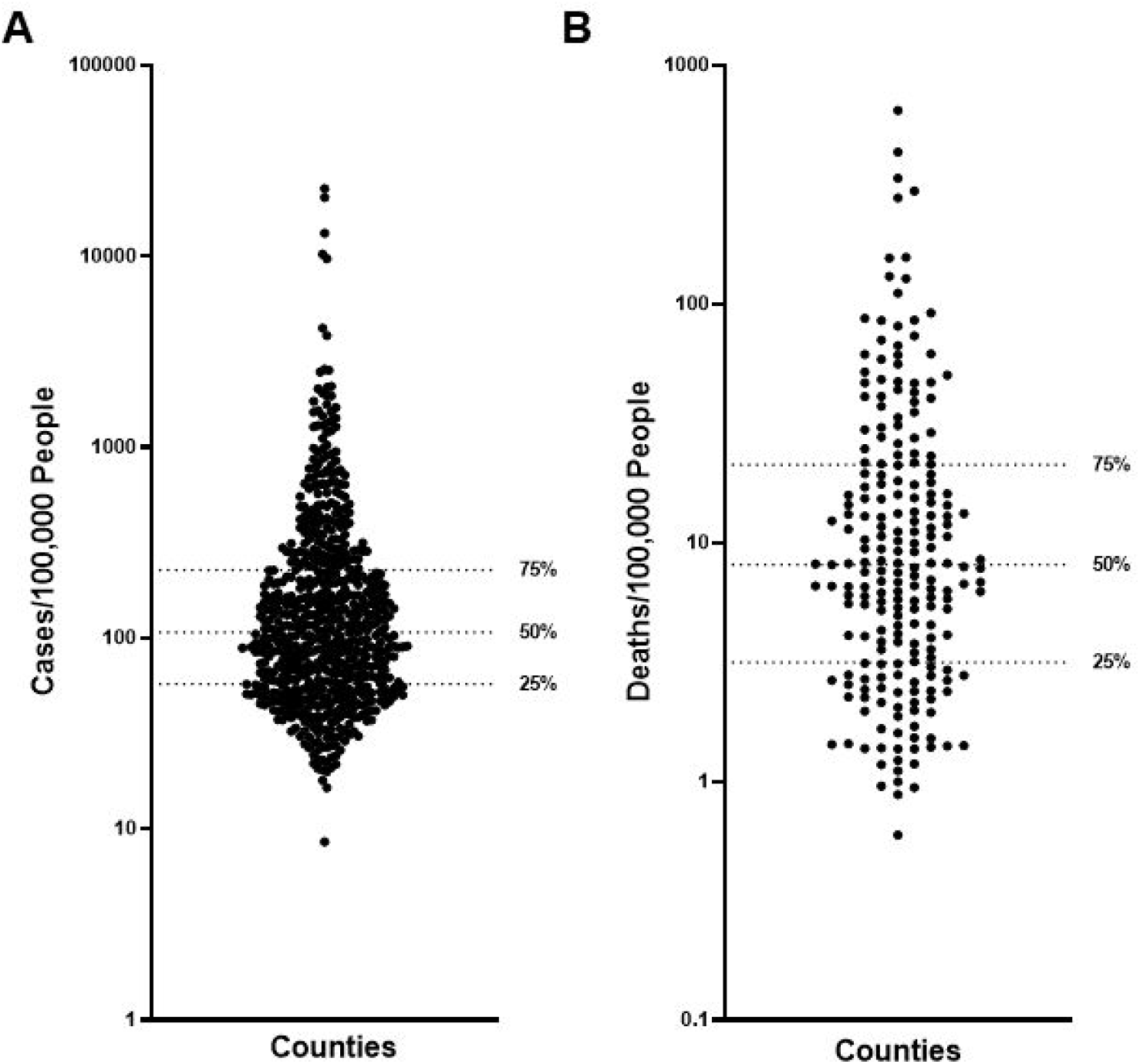
Distribution of COVID-19 **(A)** Cases/100,000 people and **(B)** Deaths/100,000 people for U.S. Counties included in analyses

County macroeconomic and COVID-19 specific variables that differed significantly between lower and upper quartiles for cases/100,000 people analysis included COVID-19 deaths/100,000 people (1.21 vs. 36.35, *P* =<0.0001), COVID-19 tests/100,000 people (832.6 vs. 1,363.3, *P* =<0.0001), GDP/Capita (45.97 vs. 85.59, *P* =0.005), percent decrease in mobility (increase in social distancing) since first COVID-19 case (42.13% vs. 38.70%, *P* =0.005), and the percent of the county living in overcrowded housing (2.94% vs. 2.42%, *P* =0.028). Nonsignificant variables in lower and upper quartile analysis included population density (594.3 vs. 1,300.6, *P* =0.116) and the number of days elapsed since the first case in the county (31.56 vs. 32.1, *P* =0.558) (**Table 3**).

**Table 3:** Characteristics of Study Cohorts Used In COVID-19 Analysis Up to April 14, 2020. ^a^ Not included in either regression model ^b^ Only included in mortality model

| Variable | Cases/100,000<br>Lowest Quartile<br>(SEM) n = 165 | Cases/100,000<br>Highest Quartile<br>(SEM) n = 165 | P Value | Deaths/100,000 Lower<br>Half (SEM)<br>n = 108 | Deaths/100,000<br>Upper Half (SEM)<br>n = 109 | P Value |
| --- | --- | --- | --- | --- | --- | --- |
| **County Macroeconomics + Covid-19 Specific Variables** |  |  |  |  |  |  |
| COVID-19<br>Cases/100,000<br>people<sup>b</sup> | 41.2 (0.82) | 1138.8 (212.9) | **<0.001** | 119.5 (8.97) | 1321.6 (318.3) | **<0.001** |
| COVID-19<br>Deaths/100,000<br>people<sup>a</sup> | 1.21 (0.08) | 36.35 (5.78) | **<0.001** | 3.79 (0.20) | 49.48 (8.41) | **<0.001** |
| COVID-19<br>Tests/100,000 people | 832.6 (27.5) | 1363.3 (58.1) | **<0.001** | 1014.1 (56.8) | 1447.7 (68.5) | **<0.001** |
| Population<br>Density/Square Mile<br>(2010) | 594.3 (67.9) | 1300.6 (444.3) | 0.12 | 1211.65 (167.81) | 1893.13 (668.46) | 0.32 |
| GDP/Capita (2010) | 45.97 (1.21) | 85.59 (13.98) | **0.005** | 61.40 (2.43) | 70.79 (11.82) | 0.44 |
| Social Distancing: %<br>Decrease in Mobility<br>After First COVID-19<br>Case | 42.13% (0.50%) | 38.70 (1.11%) | **0.005** | 43.97 (0.63) | 43.31 (1.08) | 0.60 |
| % Overcrowded<br>Housing | 2.94% (0.19%) | 2.42%(0.13%) | **0.028** | 3.06% (0.21%) | 2.37% (0.14%) | **0.007** |
| # Days Since First<br>Case | 31.56 (0.65) | 32.1 (0.57) | 0.56 | 22.14 (0.49) | 22.78 (0.57) | 0.39 |
| **County Demographics + Environmental Variables** |  |  |  |  |  |  |
| % Age >65 | 16.44% (0.35%) | 17.80% (0.31%) | **0.004** | 15.44% (0.38%) | 16.64% (0.33%) | **0.02** |
| % Non-Hispanic<br>White<sup>a</sup> | 66.64% (1.56%) | 68.01% (1.51%) | 0.53 | 58.00% (1.75%) | 65.94% (1.73%) | **0.001** |
| % Black | 9.23% (0.72%) | 15.66% (1.29%) | **<0.001** | 14.79% (1.23%) | 16.16% (1.53%) | 0.49 |
| % Hispanic<sup>a</sup> | 16.37% (1.43%) | 10.57% (0.89%) | **<0.001** | 17.83% (1.45%) | 11.16% (0.92%) | **<0.001** |
| % Asian<sup>a</sup> | 4.50% (0.44%) | 2.96% (0.30%) | **0.004** | 6.61% (0.66%) | 4.32% (0.43%) | **0.004** |
| % Native<br>Hawaiian/Other<br>Pacific Islander<sup>a</sup> | 0.34% (0.089%) | 0.11%(0.0095%) | **0.004** | 0.20% (0.028%) | 0.13% (0.015%) | **0.019** |
| % American Indian &<br>Alaska Native<sup>a</sup> | 1.14% (0.102%) | 1.61% (0.56%) | 0.41 | 1.00% (0.11%) | 1.016% (0.26%) | 0.97 |
| % Female | 50.81% (0.072%) | 50.75% (0.14%) | 0.71 | 51.02% (0.076%) | 51.10% (0.093%) | 0.52 |
| % Rural | 17.59% (1.05%) | 36.32% (2.47%) | **<0.001** | 8.07% (0.83%) | 21.90% (2.27%) | **<0.001** |
| <37° Latitude<sup>a</sup> | 49.1% (3.9%) | 31.5% (3.6%) | **0.001** | 49.1%(4.8%) | 23.9% (4.1%) | **<0.001** |
| Food Environment<br>Index | 7.73 (0.054) | 7.86 (0.092) | 0.22 | 7.756 (0.077) | 7.87 (0.102) | 0.37 |
| Violent<br>Crimes/100,000<br>People | 359.40 (16.57) | 341.59 (21.06) | 0.51 | 436.23 (22.13) | 361.13 (26.64) | **0.03** |
| Average Temperature From 10 Days Before First Case (°C) | 11.52 (0.56) | 11.33 (0.48) | 0.80 | 11.88 (0.59) | 10.54 (0.61) | 0.09 |
| Air quality: Average Ambient PM2.5 (2014) | 9.94 (0.15) | 9.37 (0.14) | **0.005** | 10.12 (0.19) | 9.76 (0.15) | 0.146 |
| % In Fair/Poor Health | 16.38% (0.292%) | 17.33% (0.33%) | **0.03** | 16.05% (0.32%) | 16.73% (0.39%) | 0.18 |
| % Poverty | 13.29% (0.37%) | 14.91% (0.53%) | **0.012** | 13.34% (0.39%) | 14.29% (0.62%) | 0.20 |
| **Medical Comorbidities + Access to Health Care** |  |  |  |  |  |  |
| % Diabetes Mellitus | 9.36% (0.15%) | 10.05% (0.31%) | **0.045** | 8.73% (0.17%) | 9.49% (0.27%) | **0.018** |
| Liver Disease Mortality/100,000 People (1998-2018) | 14.77 (0.38) | 14.95 (0.49) | 0.77 | 13.48 (0.35) | 13.68 (0.35) | 0.69 |
| Hypertension Mortality/100,000 People (2014-2016) | 233.4 (6.3) | 218.7 (8.8) | 0.18 | 223.2 (7.3) | 209.3 (8.7) | 0.22 |
| Coronary Heart Disease Mortality/100,000 People (2014-2016) | 89.14 (1.75) | 96.58 (2.04) | **0.006** | 86.87 (1.89) | 97.54 (2.51) | **<0.001** |
| Chronic Respiratory Disease Mortality/100,000 People (2014) | 56.32 (0.99) | 54.28 (1.29) | 0.80 | 50.53 (1.10) | 52.21 (1.37) | 0.34 |
| % Obesity in Ages 20+ (2015) | 29.42% (0.35%) | 29.90% (0.48%) | 0.42 | 27.37% (0.46%) | 29.34% (0.53%) | **0.005** |
| % Physical Inactivity | 22.90% (0.38%) | 25.91% (0.44%) | **<0.001** | 21.73% (0.46%) | 25.01% (0.48%) | **<0.001** |
| % Excessive Drinking | 18.99% (0.21%) | 18.21% (0.23%) | 0.01 | 19.06% (0.25%) | 18.85% (0.26) | 0.57 |
| % Smoking Tobacco | 15.54% (0.23%) | 16.72% (0.27%) | **<0.001** | 14.87% (0.29%) | 16.29% (0.36%) | **0.002** |
| Patient:Primary Care Physician Ratio | 1553.39 (58.16) | 2444.53 (232.0) | **<0.001** | 1284.96 (46.95) | 1729.78 (104.18) | **<0.001** |
| % Uninsured | 10.07% (0.36%) | 9.25% (0.29%) | 0.08 | 9.89% (0.43%) | 8.44% (0.35%) | **0.010** |
| % Flu Vaccine | 47.98% (0.46%) | 46.39% (0.56%) | **0.03** | 47.85% (0.56%) | 47.60% (0.62%) | 0.76 |

County demographic and environmental variables that were significantly different between lower and upper quartiles for cases/100,000 people analysis included percent of population over age 65 (16.44% vs. 17.80%, *P* =0.004), percent Black (9.23% vs. 15.66%, *P* =<0.0001), percent Hispanic (16.37% vs. 10.57%, *P* =0.0007), percent Asian (4.50% vs. 2.96%, *P* =0.004), percent rural (17.59% vs. 36.32%, *P* =<0.0001), average ambient PM2.5 (9.94 vs. 9.37, *P* =0.0049), percent of the population considered to be in fair or poor health (16.38% vs. 17.33%, *P* =0.033), percent living below the poverty line (13.29% vs. 14.91%, *P* =0.0124). Nonsignificant variables in lower and upper quartile analysis included percent Non-Hispanic White (66.64% vs. 68.01%, *P* =0.53), percent female (50.81% vs. 50.75%, *P* =0.71), Food Environment Index score (7.73 vs. 7.86, *P* =0.223), number of violent crimes/100,000 People (359.40 vs. 341.59, *P* =0.506), and mean daily temperature from 10 days before first case to April 14th 2020 (°C) (11.52 vs. 11.33, *P* =0.795) (**Table 3**).

County medical comorbidities and access to health care variables found to be significantly different between lower and upper quartiles for cases/100,000 people analysis included percent of population with diabetes mellitus (9.36% vs. 10.05%, *P* =0.0448), coronary heart disease mortality/100,000 (89.14 vs. 96.58, *P* =0.0060), percent considered physically inactive (22.90% vs. 25.91%, *P* =<0.0001), percent who drink excessively (18.99% vs. 18.21%, *P* =0.0125), percent who smoke tobacco (15.54% vs. 16.72%, *P* =0.0001), the patient to primary care physician ratio (1,553.4 vs. 2,444.53, *P* =0.0002), and percent who received the flu vaccine (47.98% vs. 46.39%, *P* =0.0295). Nonsignificant variables in lower and upper quartile analysis included liver disease mortality/100,000 (14.77 vs. 14.95, *P* =0.771), hypertension mortality/100,000 (233.4 vs. 218.7, *P* =0.177), chronic respiratory disease mortality/100,000 (56.32 vs. 54.28, *P* =0.804), percent considered obese for those over age 20 (29.42% vs. 29.90%, *P* =0.42), and the percent without health insurance (10.07% vs. 9.25%, *P* =0.078) (**Table 3**).

County macroeconomic and COVID-19 specific variables that were significantly different between lower and upper halves for deaths/100,000 people included COVID-19 cases/100,000 people (119.5 vs. 1,321.6, *P* =0.0002), COVID-19 tests/100,000 people (1,014.1 vs. 1,447.7, *P* =<0.0001), and percent of the population living in overcrowded housing (2.94% vs. 2.42%, *P* =0.028). Nonsignificant variables included population density (1,211.65 vs. 1,893.13, *P* =0.324), GDP/Capita (61.40 vs. 70.79, *P* =0.439), percent decrease in mobility (increase in social distancing) since first COVID-19 case (43.97% vs. 43.31%, *P* =0.604), and number of days elapsed since the first case in the county (22.14 vs. 22.78, *P* =0.393) (**Table 3**).

County demographic and environmental variables that differed significantly between lower and upper halves for deaths/100,000 people included percent of population over age 65 (15.44% vs. 16.64%, *P* =0.017), percent Non-Hispanic White (58.00% vs. 65.94%, *P* =0.0014), percent Hispanic (17.83% vs. 11.16%, *P* =0.0001), percent Asian (6.61% vs. 4.32%, *P* =0.0041), percent rural (8.07% vs. 21.90%, *P* =<0.0001), and number of violent Crimes/100,000 people (436.23 vs. 361.13, *P* =0.0311), Nonsignificant variables in lower and upper halves analysis included percent Black (14.79% vs. 16.64%, *P* =0.487), percent female (51.02% vs. 51.10%, *P* =0.5165), Food Environment Index score (7.756 vs. 7.87, *P* =0.367), mean daily temperature from 10 days before first case to April 14th 2020 (°C) (11.88 vs. 10.54, *P* =0.093), 2014 Average Ambient PM2.5 (10.12 vs. 9.76, *P* =0.1461), percent of population considered to be in fair or poor health (16.05% vs. 16.73%, *P* =0.18), and percent living below the poverty line (13.34% vs. 14.29%, *P* =0.197) (**Table 3**).

County medical comorbidities and access to health care variables that were significantly different between lower and upper halves for deaths/100,000 people included percent with diabetes mellitus (8.73% vs. 9.49%, *P* =0.018), coronary heart disease mortality/100,000 (86.87 vs. 97.54, *P* =0.0008), percent obese in those over age 20 (27.37% vs. 29.34%, *P* =0.0054), percent considered physically inactive (21.73% vs. 25.01%, *P* =<0.0001), percent who smoke tobacco (14.87% vs. 16.29%, *P* =0.0023), the patient to primary care physician ratio (1,284.96 vs. 1,729.78, *P* =0.0001), and the percent without health insurance (9.89% vs. 8.44%, *P* =0.0099). Nonsignificant variables in lower and upper halves analysis included liver disease mortality/100,000 (13.48 vs. 13.68, *P* =0.693), hypertension mortality/100,000 (223.2 vs. 209.3, *P* =0.224), chronic respiratory disease mortality/100,000 (50.53 vs. 52.21, *P* =0.342), percent who drink excessively (19.06% vs. 18.85%, *P* =0.568), and percent who received the flu vaccine (47.85% vs. 47.60%, *P* =0.760) (**Table 3**).

To control for possible confounding variables, sequential multivariate regression analyses were performed. For case rate logistic regression analysis, the adverse effect of percent Black remained significant with the addition of macroeconomic and COVID-19 specific variables in logistic regression (OR=1.03, 95% CI:1.01-1.06, *P* =<0.0001) whereas the effect of temperature was still not significant (OR=0.97, 95% CI: 0.93-1.01, *P* =0.19). Adding county demographics and environmental factors resulted in both percent Black (OR=1.16, 95% CI: 1.08-1.24, *P* =<0.0001) and temperature (OR=0.82, 95% CI: 0.73-0.90, *P* =0.0002) demonstrating significant effects on cases per 100,000. In the final model, the adverse effect of percent Black (OR=1.22, 95% CI: 1.09-1.40, *P* =0.001) and the protective effect of temperature (OR=0.81, 95% CI: 0.71-0.91, *P* =0.0009) remained robust to the addition of medical comorbidities and access to health care. Similar results are seen in the linear model, although the effects of percent Black and temperature do not become significant until Model 3 **(Table 4)**.

**Table 4:**
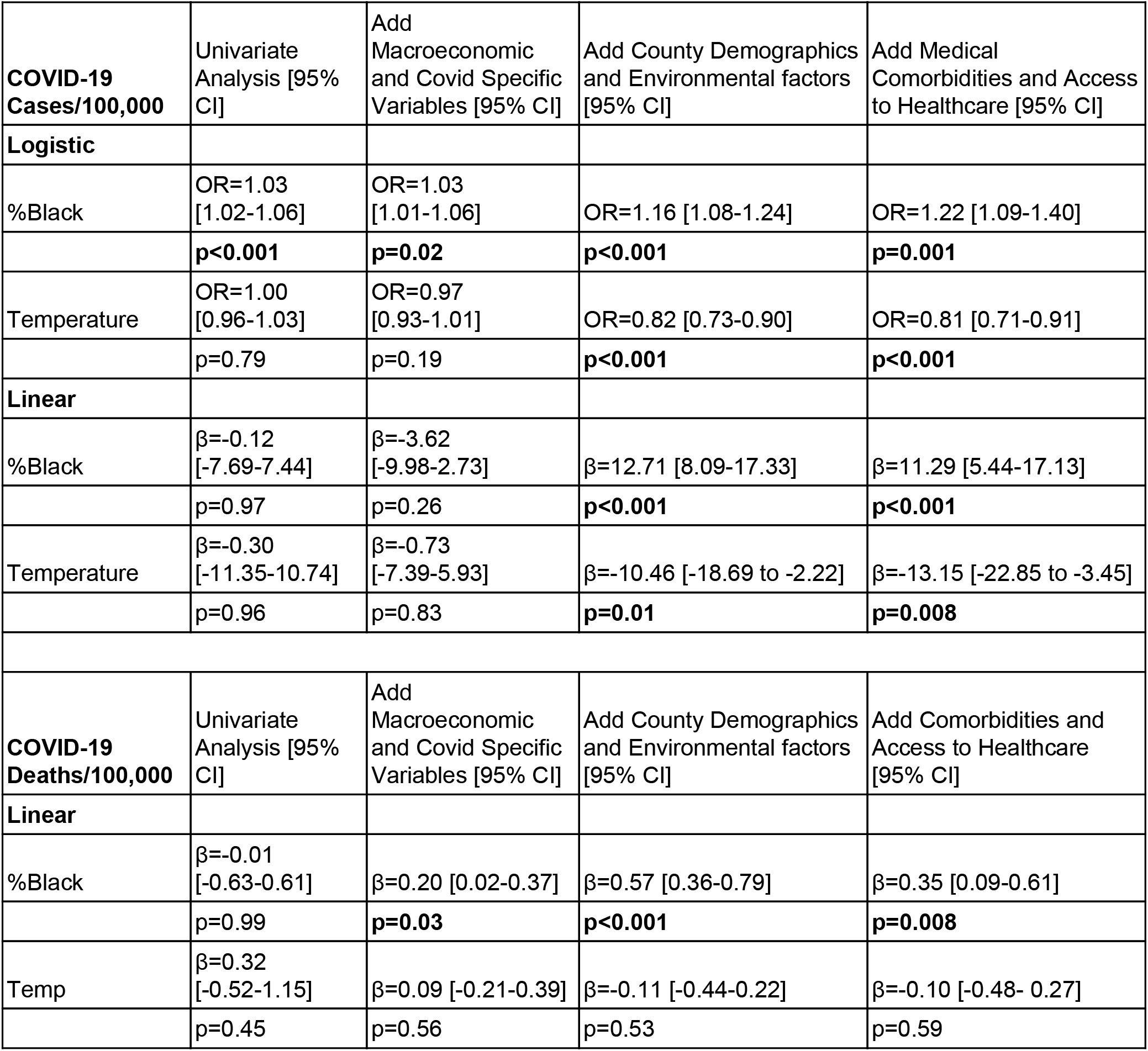
Sequential Multivariate Modeling for COVID-19 Case and Death Rate vs. Black Race and Temperature. **Top**: Logistic regression results for COVID-19 Cases per 100,000 for the lowest and highest quartiles as well as linear regression results for all 661 counties meeting the inclusion requirements. **Bottom**: Linear regression results for COVID-19 Deaths per 100,000 for all 217 counties included in the analyses. Odds ratios and their 95% confidence intervals were reported for logistic regressions. Regression coefficients and their 95% confidence intervals were reported for linear regressions

For death rate analysis, percent Black had a significantly positive effect on mortality after addition of macroeconomic and COVID-19 specific variables in the linear regression (β=0.20, 95% CI: 0.02-0.37, *P* =0.027). The effect of temperature was not significant after addition of macroeconomic and COVID-19 specific variables (β=0.09, 95% CI:-0.21-0.39, *P* =0.557). After county demographics and environmental factors were also added, the effect of percent Black (β=0.57, 95% CI: 0.36-0.79, *P* =<0.0001) remained significant while temperature was not significant (β=-0.11, 95% CI: −0.44-0.22, *P* =0.525). In the final model, after the addition of medical comorbidities and access to health care markers, the effect of percent Black (β=0.35, 95% CI: 0.09-0.61, *P* =0.008) was still significant and positive while the effect of temperature was not significant (β=-0.10, 95% CI:-0.48 - 0.27, *P* =0.594) **(Table 4)**.

## Discussion

Since late January, when the first confirmed case of COVID-19 in the United States was reported in Washington state, the U.S. has become a major epicenter of the coronavirus pandemic, now reporting the greatest number of total cases and deaths worldwide. Recent work has primarily focused on patient demographics, underlying health comorbidities, social disparities in healthcare access and quality, and environmental variables such as pollution to identify potential risk factors and vulnerable populations.^6-20^ However, while these previous studies have examined the effects of these domains on COVID-19 spread independently, some do not control for the potentially confounding interactions between variables. In this study, we sought to investigate COVID-19 prevalence and mortality in all U.S. counties through a more comprehensive framework that accounts for effects of county-level macroeconomic, demographic, environmental, health status, and healthcare access variables. By conducting sequential multivariate analyses of variables that span social, structural, and environmental spheres, our study aims to more completely characterize the epidemiology of SARS-CoV-2 and thereby identify particularly salient risk factors and vulnerable populations in the United States.

### Impact of Race

In recent weeks, headlines and research studies have brought race to the forefront as a potentially significant risk factor for increased COVID-19 incidence in the United States, revealing that African Americans have accounted for all but three COVID-19 related deaths in St. Louis, Missouri and three quarters of those in Milwaukee, Wisconsin.^31,32^ Further, recent analyses of patients hospitalized for COVID-19 in 99 U.S. counties spanning 14 states have demonstrated significantly increased hospitalization rates amongst Black Americans.^6,31,33^ Our study corroborates and expands on these findings to suggest that Black Americans may be at significantly higher risk of COVID-19 infection and mortality nationwide. However, at this time, current theories to explain this disproportionate burden of COVID-19 morbidity and mortality in the Black population have remained speculative and limited by confounding variables. Recent studies have pointed to the fact that, compared to the general population of the United States, Black Americans have been shown to suffer from increased rates of chronic medical comorbidities such as cardiovascular disease, diabetes mellitus, hypertension, obesity, and chronic respiratory disease, all of which have been shown to convey increased risk of SARS-CoV-2 infection and lead to worse outcomes.^18-20^ Studies have also cited inequities in structural variables, which manifest in the disproportionate number of Black Americans who suffer from poverty, reside in densely packed areas with more environmental hazards, have decreased access to healthy food sources, and lack healthcare coverage and access.^31^ In our comprehensive multivariate analysis, we were able to control for these potentially confounding variables; however, after doing so, our data continue to demonstrate a disproportionate number of COVID-19 cases and deaths in counties with large Black populations in the United States.

This increased disease burden did not seem to be explained by previously proposed mechanisms, indicating that other modulating factors should be considered and novel interventions designed appropriately. One potential domain includes additional sociocultural variables that have only now become relevant within the context of the pandemic. For example, African Americans make up a large percentage of the healthcare, transportation, government, and food supply industries, job sectors that have now been deemed “essential” services in light of the SARS-CoV-2 pandemic.^34^ Despite the majority of Americans currently living in counties with some form of social distancing mandate, fewer than one in five Black Americans have a job that gives them this flexibility to work from home, compared with more than a third of white and Asian American workers.^35^ The potentially fatal consequences of this fact can already be seen in the case of the New York City transit workforce, which has been among the hardest hit by the virus with more than 2,000 cases and 59 deaths.^57^ Despite making up just 24% of the city’s overall population, African Americans account for nearly half of transit workers.^36,37^ Furthermore, despite an 87% reduction in overall ridership since the pandemic began, the nearly one million remaining passengers–most of whom lack the luxury of social distancing due to essential employment–are predominantly low-income people of color.^38^

However, this theory is complicated by the fact that Hispanic workers also makeup a disproportionate percentage of the essential workforce, yet our data do not suggest an increased burden of COVID-19 cases and mortality amongst counties with greater Hispanic populations.^35^ A potential explanation may be that Black workers may be more likely than other minority groups to work essential jobs that also require close proximity and frequent contact with others, such as bus drivers, postal workers, and grocery store clerks, which would convey additional risk of COVID-19 infection.^39,40^ However, more work is needed to elucidate potential mechanisms to explain the differing disease burdens between these vulnerable populations.

Another potential consideration is that non-Hispanic Black populations in the United States have been shown to suffer from vitamin D deficiency at rates much higher than any other ethnic or racial group, with recent data showing that Black and Hispanic populations in the United States suffer from vitamin D deficiency at rates of 24 and 4 times more than those seen in the white population, respectively.^41^ In this comprehensive analysis of serum 25-hydroxyvitamin D (25(OH)D) levels collected from more than 26,000 adults in the United States between 2001-2010 as part of the National Health and Nutrition Examination Survey (NHANES), 71.9% of non-Hispanic Black individuals were found to suffer from vitamin D deficiency (defined as serum concentrations of less than 50 nmol/L) even after controlling for other potential predictors.^41^ Of the many sociodemographic, behavioral, and clinical variables studied, being a racial or ethnic minority was the strongest predictor of vitamin D deficiency.^41^

Vitamin D has been suggested as a potentially mitigative factor in the COVID-19 pandemic due to its important modulatory effect on immune response.^42,43^ Furthermore, vitamin D deficiency has been implicated in numerous adverse health conditions such as acute respiratory syndromes, tuberculosis, cardiovascular disease, autoimmune disease, and some cancers.^44^ This higher prevalence of vitamin D deficiency in darker-skinned individuals is thought to largely be due to increased melanin pigmentation, which absorbs significantly more radiation from sunlight and thereby reduces the available ultraviolet-B radiation that is needed to trigger natural vitamin D production in the skin.^45^

### Impact of Temperature

The potential role of vitamin D as a modulator of COVID-19 burden is further supported by our other significant finding regarding temperature, which suggests a protective effect of warmer climates. Several recent reports have also linked higher temperature to decreased virus spread, while others have found no significant effects.^9-14^ Here, we demonstrate an independent effect of temperature that results in reduced COVID-19 cases, but not mortality, across U.S. counties. If warmer temperatures do, in fact, play a role in mitigating disease spread, it is reasonable to expect a potential seasonal trend in global cases and mortality. As temperatures begin to warm in the Northern hemisphere during the summer months, we may begin to see a decrease in disease burden; however, we should prepare to see a resurgence in COVID-19 incidence when temperatures again decrease in the fall and winter months, as has been exhibited in previous pandemics like the Spanish flu.^46^ Likewise, countries in the Southern hemisphere should be prepared to experience seasonal trends in the opposite direction.

The mechanism through which temperature may execute its mitigative effects remains unclear, though it has already been posited that increased vitamin D levels due to greater sun exposure may result in better immune response against the SARS-CoV-2 virus, as it has been shown to do within the context of other viruses.^42,43^ Notably, when temperature is substituted out of our final model for other vitamin D proxies, such as average daily sunlight and latitude above and below 37 degrees, the results are also highly significant in the same direction. Including these variables together in the model resulted in multicollinearity, which suggests that they are most likely measuring the same effect. Thus, in this study, average temperature prior to the first reported COVID-19 case was chosen to be the vitamin D proxy because it was the continuous variable that contained the most specific and validated data at the county level.

One potential clinical implication of these findings is on the use of chloroquine and hydroxychloroquine therapy in treatment of the SARS-CoV-2 virus, which has shown some promise in early research.^47^ Chloroquine has been shown to reduce the conversion of vitamin D from calcidiol to calcitriol, the former of which has been demonstrated to potentiate the immune system,^48^ which may serve as a partial mechanism for the purported beneficial effects of chloroquine therapy on COVID-19 outcomes. Future research on the role of serum vitamin D levels on COVID-19 outcomes are already underway, including a retrospective review of patients with documented vitamin D levels at our institution, as well as a prospective randomized, controlled trial at the University of Grenada that is investigating the role of vitamin D supplementation therapy versus placebo on outcomes in 200 COVID-19 patients.^49^

Our results support the hypothesis that vitamin D supplementation, whether through behavioral or dietary interventions, may prove to be beneficial for SARS-CoV-2 outcomes in the United States. Though the potential benefits of vitamin D supplementation seem to be particularly relevant for racial and ethnic minorities, they should also be considered for the entire population, as studies have found that less than 30% of U.S. adults have vitamin D levels considered sufficient for optimal health outcomes.^41^ Though standardized data on worldwide vitamin D levels are currently lacking, existing meta-analyses suggest that vitamin D deficiency is also a rampant issue on a global scale and should be prioritized for future research.^50^ If the theorized effects of vitamin D on COVID-19 morbidity are supported by ongoing studies, vitamin D deficiency could emerge as a unifying theory for the findings herein and allow countries to mobilize resources in anticipation of seasonal trends and develop targeted interventions to mitigate risk.

### Additional Factors

Other findings associated with increased COVID-19 cases and deaths included increased poverty, increased GDP per capita, increased pollution, and increased flu vaccination rate. Poverty and GDP are not typically thought to be positively correlated with each other, however, the correlations seen here may represent inequalities in health care versus and access to testing.For example, poverty may exacerbate case spread, while economic power increases testing rates, both of which would lead to increased cases.^47^ Flu vaccination was included as a marker for access to healthcare and may be associated with greater testing rates. Additionally, flu vaccination is often mandated for healthcare workers, thus counties with higher flu vaccination rates may have more of its citizens having front line exposure to the virus.^51,52^

Decreased pollution was associated with increased COVID-19 cases, which does not align with prior findings in studies that associated increased air pollution with increased mortality.^15,29^ Our model also shows an inconclusive connection between air quality and COVID-19 deaths. However, air pollution data in this study is from 2014 and may not reflect current air pollution. Given increased social distancing, many counties with historically bad air pollution currently have significantly decreased pollution levels, which may have decreased pollution related COVID-19 cases.^53,54^

Although the effect was occasionally significant in some steps of the sequential regression models, increased social distancing as measured by the average reduction in cellphone movement from the time of the first case to the end of the study was not a robust modulator of cases or deaths (data not shown). The most likely reason for this is due to the simultaneity bias of this metric. In particular, the metric used here does not differentiate between proactive counties and reactive counties. It is expected that less proactive social distancing will cause more cases and deaths (negative correlation) while higher numbers of cases and deaths induces fear, causing increased reactive social distancing (positive correlation). Since these effects work in opposite directions, this bias makes it more difficult to find an effect of social distancing. Studies that plan to model social distancing with numerous covariates should ensure their metric adequately delineates between proactive and reactive social distancing or employ an appropriate instrument to remove this simultaneity.^55^

### Limitations

The emerging nature of Covid-19 required inclusion of only counties with sufficient data. Many counties were excluded from analysis for lack of Covid-19 cases or lack of Covid-19 deaths. This limitation is unavoidable and waiting for sufficient data may deter public health response. This study is also limited by the fact the COVID-19 pandemic is still in progress in all of the counties included here and is at different stages. Thus, the number of cases and mortality statistics for the counties provides a snapshot of the current state, but may not, and likely will not, be reflective of the ultimate case and death tolls in these counties. Moreover, most counties are still in the early stages of accumulating deaths which axiomatically lags behind the number of cases. While our results of temperature and Black race on cases per 100,000 have been consistent for a number of weeks, the number of counties with at least 10 deaths has only recently become large enough to perform similar analyses. Temperature also demonstrates this lack of simultaneity, but changes more predictably and slowly than Covid-19 cases and deaths. Additionally, since we relied heavily on publicly available and easily accessible sources to create our database, we used sources that may be outdated or otherwise inconsistent with the actual value of various statistics during the past few months. Confounding effects are also possible, given that it is impossible to fully extricate socioeconomic, demographic, and even some environmental variables from each other. The stepwise approach to regression mitigates excessive confounding risk through sensitivity analysis and signifies that the findings are not error artifacts. Ecological fallacies may be present in the model for demographic and socioeconomic variables compared to individual cases and deaths; we report as granularly as practical to account for this.

## Conclusion

This study evaluated the independent effects of Black race and temperature on the incidence of cases and mortality of COVID-19 at the U.S. county level. In multivariate regression analyses that controlled for county demographics, socioeconomic factors, and medical comorbidities, counties with higher average daily temperatures were associated with decreased COVID-19 cases, but not deaths. Black race was significantly associated with both increased cases and increased deaths. This suggests that many of the proposed mechanisms through which Black race might increase risk for COVID-19 such as socioeconomic and healthcare-related predispositions, are inadequate in explaining the full magnitude of this health disparity. A potential unifying theory of these results is the preponderance of vitamin D deficiency of Black citizens in comparison to other races in the U.S. However, additional study is needed to further understand these results.

## Data Availability

Data is publicly available on GitHub.

https://github.com/johndurbin93/Covid-19-Dataset

## Acknowledgments

We thank Lauren Spinazze from the Unacast Group for providing us with data from their Social Distancing Scoreboard.^24^ We would also like to thank Aman M. Choudhri for his help with data acquisition.

## Notes

### Competing Interest Statement

The authors have declared no competing interest.

### Funding Statement

No external funding was received

